# Resting energy expenditure and thermic effect of a high-fat meal in the early follicular and mid-luteal phases of the menstrual cycle: a crossover trial protocol

**DOI:** 10.64898/2026.05.25.26354032

**Authors:** Nicholas Goulet, Sydney Lyndon, Noémie Beauregard, Kurt McInnis, Jean-François Mauger, Éric Doucet, Pascal Imbeault

## Abstract

**Introduction:** Menstrual cycle phase has been proposed as a source of intra-individual variability in resting energy expenditure and the thermic effect of food in premenopausal females, yet studies examining the thermic effect of food across menstrual cycle phases report conflicting findings.

**Methods:** This protocol describes a secondary analysis of prespecified outcomes from a non-randomized, two-period crossover trial primarily designed to assess postprandial plasma triglyceride concentrations across menstrual cycle phases (ClinicalTrials.gov: NCT07459465) in 12 premenopausal females aged 18–30 years, free of chronic disease and hormonal contraceptive use, recruited in Ottawa, Canada. Participants will complete two experimental sessions: one in the early follicular phase and one in the mid-luteal phase, each involving consumption of a high-fat meal. Eleven secondary outcomes will be reported: fasting resting energy expenditure, thermic effect of food, respiratory exchange ratio, carbohydrate oxidation rate, lipid oxidation rate, desire to eat, hunger, fullness, prospective food consumption, serum β-estradiol, and serum progesterone. Masked outcome analyses will be performed using linear mixed-effects models.

**Results:** Recruitment began on 26 March 2026; results will be reported in the Stage 2 manuscript.

**Discussion:** Findings from this trial may help clarify whether menstrual cycle phase constitutes a meaningful source of intra-individual variability in energy metabolism, with implications for the design of metabolic research in premenopausal females.

## INTRODUCTION

Resting energy expenditure and the thermic effect of food are two key components of total energy expenditure, together accounting for the majority of an individual’s daily energy needs in sedentary to lightly active individuals (1,2). In this context, resting energy expenditure, defined as the energy required to sustain basic physiological functions at rest, constitutes approximately 60– 70 % of total energy expenditure, while the thermic effect of food, reflecting the metabolic cost of digesting, absorbing, transporting, and storing nutrients, contributes a further ∼10 %, with physical activity accounting for the remainder (3–5). Day-to-day intra-individual variation in premenopausal females, including variations in reproductive hormones, has been proposed as a potential modulator of both fasting resting energy expenditure and the thermic effect of food (6,7). In premenopausal females, circulating concentrations of reproductive hormones fluctuate substantially across the menstrual cycle: estradiol varies approximately fivefold and progesterone more than 50-fold between the early follicular and mid-luteal phases (8,9). However, whether these cyclical hormonal fluctuations constitute a meaningful intra-individual source of variability in fasting resting energy expenditure and the thermic effect of food remains poorly understood.

The potential influence of menstrual cycle phases on fasting resting energy expenditure is further supported by a systematic review and meta-analysis of 26 studies involving 318 premenopausal females, which reported a small but statistically significant increase (standardized mean difference = 0.33; 95 % confidence interval: 0.17–0.49) in resting energy expenditure during the luteal phase compared with the follicular phase (10). However, most included studies were reportedly of low methodological quality, with inadequate statistical power, poor control of potential confounders (e.g., environmental temperature, physical activity, and dietary and substance use before testing), and relied on calendar counting or basal body temperature rather than biochemical verification of menstrual cycle phases (10). To our knowledge, only two studies have examined the effect of menstrual cycle phases on the thermic effect of food. Piers et al. reported an 18.5 % increase in the thermic effect of a standard test meal (1880 kJ; 75 % carbohydrate, 15 % fat, 10 % protein) over 300 minutes of measurement during the luteal phase relative to the follicular phase in 13 females, with phase confirmed by plasma concentrations of estradiol and progesterone (11). In contrast, Tai et al. observed a 19 % reduction in the thermic effect of a high-fat liquid meal (3138 kJ; 54.5 % carbohydrate, 31.5 % fat, 14.0 % protein) over 205 minutes of measurement during the luteal phase compared with the early follicular phase in eight females, using basal body temperature to estimate phase (12). Notably, the two studies also differed in meal composition, total energy content, postprandial measurement duration, and study population.

Beyond phase comparisons, evidence from studies examining hormonal status more broadly also suggests that reproductive hormones may modulate the thermogenic response to a meal. Duhita et al. demonstrated that females using a combined monophasic oral contraceptive pill showed no significant difference in the thermic effect of food across a threefold range of meal protein content, a thermogenic inflexibility not observed in females without oral contraceptive use, who exhibited a significant dose-dependent increase with higher protein intake (13). More recently, a secondary analysis of 86 females approaching menopause found that habitual protein intake was the only statistically significant predictor of the thermic effect of food, explaining just 6 % of its variance, while follicle-stimulating hormone offered no additional explanatory value, alongside body composition and fitness (7). The direction, magnitude, and hormonal basis of any menstrual cycle effect on fasting resting energy expenditure and the thermic effect of food, therefore, remain unclear.

This study will examine whether fasting resting energy expenditure and the thermic effect of a high-fat meal over 6 hours differ between the early follicular (low circulating ovarian sex steroid hormone concentrations) and mid-luteal (high circulating ovarian sex steroid hormone concentrations) phases of the menstrual cycle. Specifically, we will report 11 secondary outcomes from a crossover trial primarily designed to characterize postprandial plasma triglyceride concentrations across menstrual cycle phases in premenopausal females; the primary results will be reported in a separate manuscript. Secondary outcomes include fasting resting energy expenditure, thermic effect of food, respiratory exchange ratio, carbohydrate and lipid oxidation rates, subjective appetite scores (desire to eat, hunger, fullness, prospective food consumption), and serum β-estradiol and progesterone concentrations. Given the conflicting findings from prior studies and the exploratory nature of this secondary analysis, no formal a priori hypothesis regarding the direction of any menstrual cycle phase effect was developed.

## METHODS

This registered report constitutes a Level 4 submission per the Peer Community In Registered Reports taxonomy. Some of the data to be used in the planned analyses have already been acquired and are in the authors’ possession; however, the authors hereby self-certify that none of the data relevant to the present secondary outcomes, nor the primary outcomes, have been analyzed prior to submission of this Stage 1 protocol.

### Research question

Does menstrual cycle phase influence the thermic effect of food following consumption of a high-fat meal in premenopausal females?

### Research ethics approval

This study was approved by the University of Ottawa Health Sciences and Science Research Ethics Board (file number: H-06-18-837) and will be conducted in accordance with the Declaration of Helsinki. All participants will provide written informed consent prior to enrolment.

### Patient and public involvement

Patients and members of the public were not and will not be involved in the design, conduct, or reporting of this trial.

### Trial design

The trial was prospectively registered on ClinicalTrials.gov (identifier: NCT07459465, registered 03 March 2026; trial incomplete and open to new participants at the time of manuscript submission; https://clinicaltrials.gov/study/NCT07459465). The trial will use a single-site, non-randomized, two-period, crossover design with a superiority framework. This report will present 11 secondary outcomes from a trial primarily designed to assess postprandial plasma triglyceride concentrations across menstrual cycle phases. Trial results for the seven primary outcomes (i.e., plasma concentrations of total triglycerides, buoyant triglycerides, denser triglycerides, non-esterified fatty acids, insulin, glucose, and β-hydroxybutyrate) will be reported in a separate manuscript. The Standard Protocol Items: Recommendations for Interventional Trials (SPIRIT) and the Consolidated Standards of Reporting Trials (CONSORT) 2025 Statements were followed in the preparation of this article to ensure complete and transparent reporting (14,15). Figure 1 contains a timeline that details the schedule and time commitment for trial participants.

**Figure 1.**
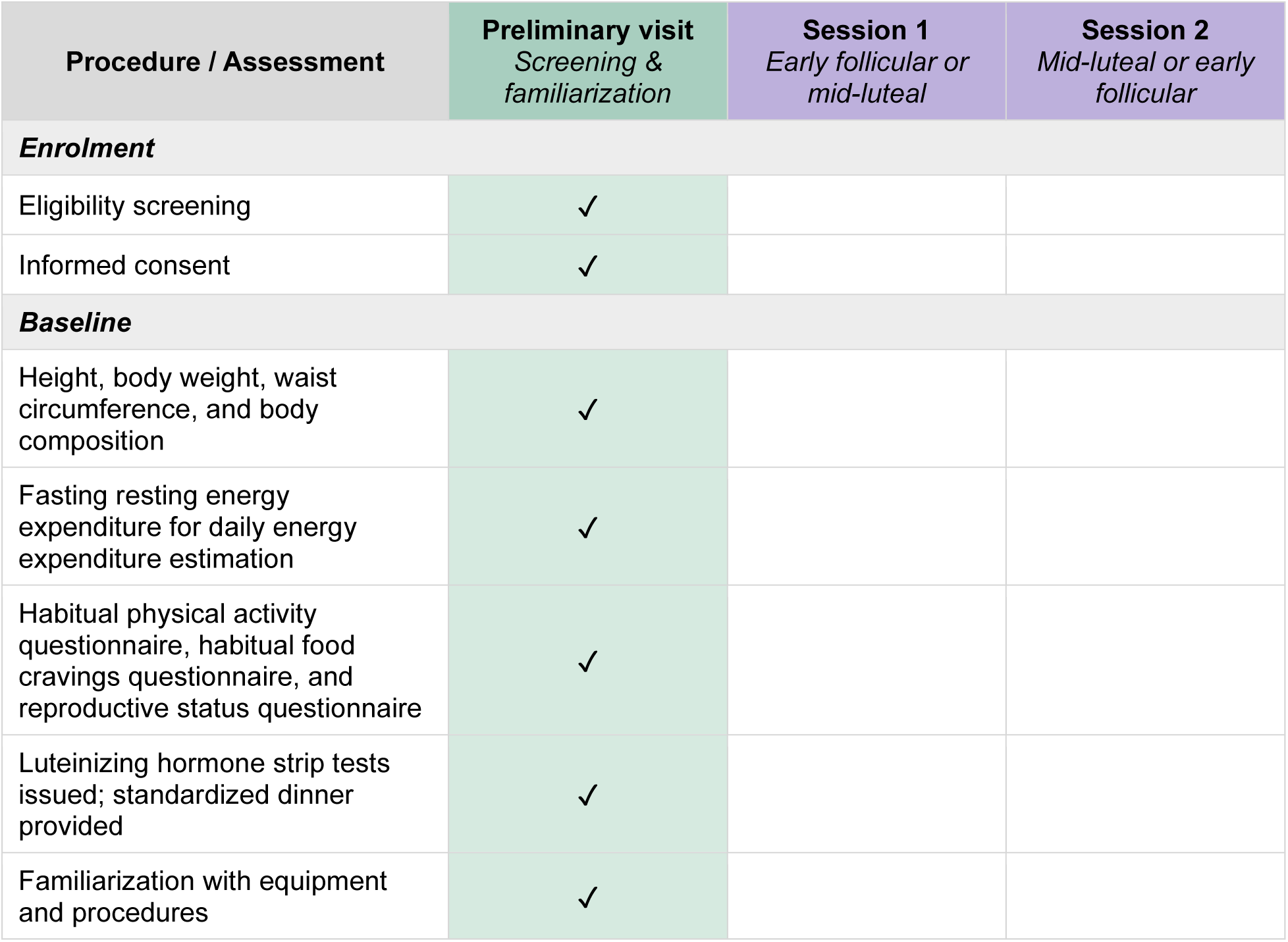
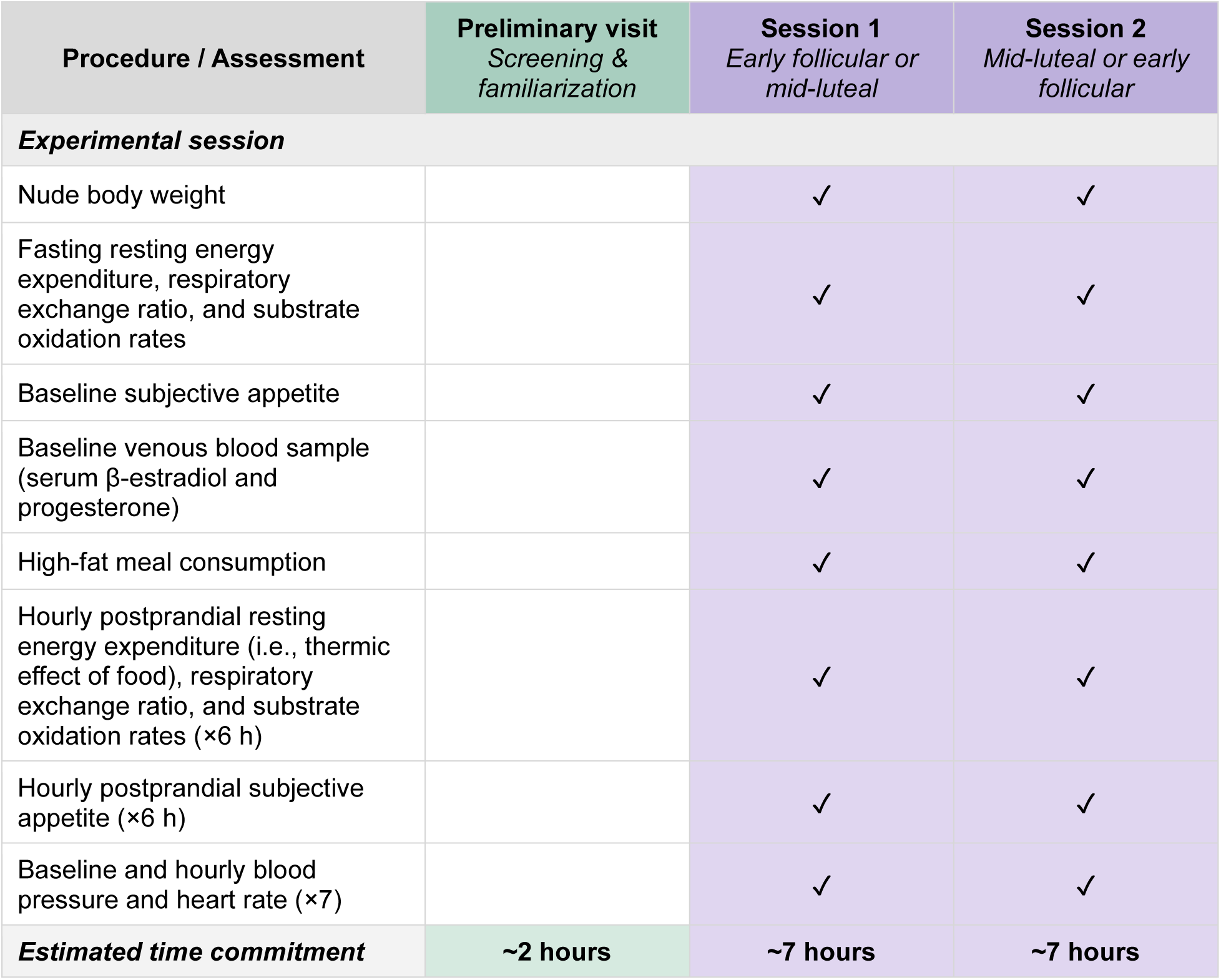
Schematic diagram outlining the schedule and time commitment for trial participants.

### Changes to trial protocol

Three secondary outcomes (i.e., fasting resting energy expenditure, carbohydrate oxidation rate, and lipid oxidation rate) were added to the trial registry on 15 May 2026, following its initial release, bringing the total to 11. No other modifications have been made to the trial protocol or registry.

### Trial setting

All experimental sessions will be conducted at the Behavioural and Metabolic Research Unit, School of Human Kinetics, Faculty of Health Sciences, University of Ottawa, Ottawa, Ontario, Canada. Participants will be recruited from the community (non-clinical setting). Recruitment began on 26 March 2026, and the expected study completion date is 31 August 2026.

### Eligibility criteria

Community-dwelling adults will be recruited from the Ottawa–Gatineau region via convenience sampling (e.g., university students). Inclusion criteria will be: age 18–30 years; female biological sex; ability to communicate in English or French; and ability to provide informed consent. Exclusion criteria will be: history or evidence of chronic disease; current use of hypolipemic medication, hormonal contraceptives, antidepressants, anticoagulants; ongoing smoking status; and experiencing pregnancy, puerperium, or irregular menstrual cycles, defined as self-reported cycle lengths outside the 21–35-day range for at least three consecutive cycles prior to enrolment.

### Preliminary screening session

All participants will attend a preliminary screening visit to the research unit, during which informed consent will be obtained, and eligibility confirmed. All screening sessions will be scheduled between 07:00 and 09:00. Body height and mass will be measured using a stadiometer (HR-100 Height Rod, Tanita Corporation of America) and a digital weighing terminal (HR-100, BWB-800AS, Tanita Corporation of America), respectively. Body mass index will be calculated as mass in kilograms divided by the square of height in meters. Waist circumference will be measured in duplicate at the level of the umbilicus in accordance with World Health Organization guidelines (16). Body composition will be assessed by dual-energy X-ray absorptiometry (Lunar Prodigy, General Electric). The coefficient of variation and correlation for repeated body composition measurements in our laboratory are 1.8% and r = 0.99, respectively, as determined in a separate sample of 12 healthy individuals (4 females). Following a 12-hour overnight fast, resting energy expenditure will be measured by indirect calorimetry with a research-grade metabolic testing system (Vmax Encore 29 N, VIASYS Sensor Medics Corporation) and a ventilated hood in a thermoneutral (∼22 °C), darkened environment, with participants resting in a semi-recumbent position for 30 minutes; the first and last 5 minutes will be excluded from analysis. Participants will undergo 10 minutes of supine rest prior to measurement of fasting energy expenditure, during which the calorimeter will be set up and calibrated. Achievement of steady state will be reported in the Stage 2 manuscript, defined as a minimum of 5 consecutive minutes during which the coefficient of variation for energy expenditure, averaged over 1-minute intervals, is less than 10%. The coefficient of variation and correlation for repeated fasting resting energy expenditure measurements in our laboratory are 1.4% and r = 0.99, respectively, as determined in a separate sample of ten healthy individuals (2 females). Daily total energy expenditure will be estimated by multiplying the measured fasting resting energy expenditure by a physical activity factor of 1.375, corresponding to light physical activity, which was selected to reflect the sedentary conditions of the experimental session (17).

Habitual physical activity level will be assessed using the Canadian Society for Exercise Physiology Get Active Questionnaire (18). Habitual food cravings will be assessed using the Food Cravings Questionnaire—Trait—reduced, a validated 15-item self-report instrument measuring the tendency to experience food cravings (19). Reproductive status will be assessed using the Reproductive Status Questionnaire for Menstrual Cycle Studies (20). Participants will be familiarized with the testing equipment and procedures at this preliminary visit. Participants will also be provided with a standardized dinner to consume 12 hours before arriving at the laboratory for their experimental sessions, consisting of either a meat lasagna (390 kcal; 43 % carbohydrate, 33 % fat, 24 % protein; President’s Choice) or a vegetarian macaroni and cheese (340 kcal; 60 % carbohydrate, 21 % fat, 19 % protein; President’s Choice), based on dietary preference and held constant for each participant. Participants will be instructed to consume this meal as their final food intake before the 12-hour overnight fast. In the 36 hours preceding each experimental session, participants will be instructed to avoid vigorous physical activity, excessive consumption of alcohol and caffeine, and use of anti-inflammatory medication. Participants will also be instructed to obtain a minimum of 7 hours of sleep the night before each session. Compliance with these instructions will be verified verbally upon arrival at the laboratory. Any deviation will be recorded and reported transparently.

### Intervention and comparator

All consented participants attend two experimental sessions. Unlike pharmacological or dietary interventions, menstrual cycle phases are naturally occurring physiological states that cannot be administered or withheld. Accordingly, this study will not involve a conventional intervention and comparator in the clinical trial sense. Experimental session order will be determined by scheduling convenience (follicular then luteal, or luteal then follicular), with both sessions ideally completed within two consecutive menstrual cycles. If sessions cannot be completed within two consecutive cycles, data will nonetheless be retained, and the inter-session interval will be reported descriptively in the Stage 2 manuscript. The early follicular phase will be defined as days 1–5 of menstruation, identified by self-reported onset of bleeding. The mid-luteal phase will be defined as 7–9 days after a positive result on a urinary luteinizing hormone strip test, provided to participants during the preliminary visit. Participants will begin luteinizing hormone testing twice daily (morning and evening) on day 5, 10, or 12 of the cycle for self-reported usual cycle lengths of 21–25, 26–30, or 31–35 days, respectively, and photograph each positive result for transmission to the research team. A confirmed positive test will be required before each mid-luteal session is scheduled. If no positive test is obtained within a given cycle, suggesting an anovulatory cycle, testing will resume in the following cycle, and the session will be rescheduled accordingly. Participants will also log the start date of each menstrual period from enrollment until their experimental sessions are complete. If menstruation occurs within the 9-day window following a positive urinary luteinizing hormone strip test, scheduled experimental sessions for the luteal phase will be rescheduled. Both menstrual cycle phases will be confirmed biochemically by analyzing serum levels of estradiol and progesterone after data collection is complete. All experimental sessions will be scheduled between 07:00 and 09:00, with exact arrival times held constant for each participant across sessions to minimize the influence of circadian variation on resting energy expenditure measurements.

Upon arrival for each session, following a 12-hour overnight fast, nude body weight will be measured, and participants will then rest in a semi-recumbent position in a thermoneutral (∼22 °C), darkened environment for 30 minutes to measure resting energy expenditure. Subjective appetite scores will be recorded before inserting an intravenous catheter into the antecubital vein and collecting a baseline blood sample, prior to consuming the high-fat meal. The high-fat meal will consist of Ensure Plus (Abbott Laboratories) and 35 % whipping cream (Neilson), with the volumes of each component calculated individually for each participant to provide exactly 33 % of their estimated daily total energy expenditure. The required volumes will be derived by solving the following system of two equations:

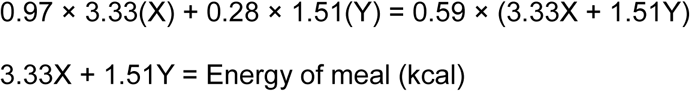

where X is the volume of whipping cream (mL), Y is the volume of Ensure Plus (mL), 3.33 kcal/mL and 1.51 kcal/mL are the energy densities of whipping cream and Ensure Plus, respectively, 0.97 and 0.28 are the proportions of energy derived from fat in whipping cream and Ensure Plus, respectively, 0.59 is the target proportion of energy from fat, and Energy of meal is the participant’s target meal energy content (kcal). As this analysis will report secondary outcomes from a trial primarily designed to assess postprandial plasma triglyceride responses, a high-fat meal was selected to maximize the primary trial’s postprandial triglyceride excursion, consistent with standard protocols in postprandial lipemia research. The resulting meal will provide 59 % of energy from fat, 31 % from carbohydrates, and 10 % from protein. Water will be available ad libitum, except when the ventilated hood is in place for indirect calorimetry; fluid intake will be recorded using an electronic balance (Scout Pro SP2001, OHAUS) and will be reported descriptively. The thermic effect of food, respiratory exchange ratio, carbohydrate and lipid oxidation rates, and subjective appetite scores will be reassessed hourly in the same thermoneutral room (∼22 °C), darkened during indirect calorimetry. Participants may withdraw at any time. Sessions will be discontinued if a participant experiences any adverse event requiring medical attention. Data from incomplete sessions will be retained, and any discontinuity will be transparently reported. Participants will be permitted to continue any medications or supplements declared at screening that do not meet the exclusion criteria.

### Outcomes

The primary outcomes of the parent trial (i.e., plasma concentrations of total triglycerides, buoyant triglycerides, denser triglycerides, non-esterified fatty acids, insulin, glucose, and β-hydroxybutyrate) will be reported in a separate manuscript. Secondary outcomes were selected to characterize the energy expenditure and appetite responses to a high-fat meal across menstrual cycle phases, and to confirm hormonal phase assignment. These outcomes are not part of a published core outcome set for this domain. The present analysis will report all 11 secondary outcomes: resting energy expenditure, thermic effect of food, respiratory exchange ratio, carbohydrate and lipid oxidation rates, desire to eat, hunger, fullness, prospective food consumption, serum β-estradiol concentrations, and serum progesterone concentrations. All data will be collected by S.L. Secondary outcomes will be assessed by N.G., N.B., and K.M.

The thermic effect of food (kcal/min) will be defined as the increase in resting energy expenditure above the fasting baseline value and will be quantified at each postprandial time point by subtracting fasting resting energy expenditure from postprandial resting energy expenditure. The total thermic effect of food over the 360-minute postprandial period will be estimated by summing the product of each hourly value and 60 minutes, assuming that each measurement represents the mean thermogenic response over the preceding hour. The thermic effect of food will also be expressed as a percentage of the test meal’s energy content. Descriptive data will be presented as means and standard deviations; figures will include individual participant data points alongside group means.

#### Energy expenditure and substrate oxidation rates

Fasting resting energy expenditure, thermic effect of food, the respiratory exchange ratio, and carbohydrate and lipid oxidation rates will be calculated from indirect calorimetry measurements with a research-grade metabolic testing system (Vmax Encore 29 N, VIASYS Sensor Medics Corporation) and a ventilated hood at baseline for 30 minutes (first and last 5 minutes excluded from analysis) and during the last 20 minutes of each hour for 6 hours following meal consumption (first and last 3 minutes excluded from analysis). Participants will undergo 10 minutes of supine rest prior to measurement of fasting energy expenditure, during which the calorimeter will be set up and calibrated. Achievement of steady state will be reported in the Stage 2 manuscript, defined as a minimum of 5 consecutive minutes during which the coefficient of variation for energy expenditure, averaged over 1-minute intervals, is less than 10%. Ethanol burn tests will be performed regularly throughout the trial to verify the accuracy of the metabolic testing system, in addition to standard gas and volume calibration performed before each session. Carbohydrate and lipid oxidation rates will be calculated using the following equations (21), assuming protein oxidation accounts for 10 % of total energy expenditure:

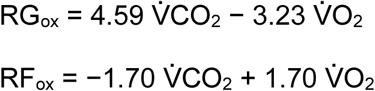

where RG_ox_ and RF_ox_ denote carbohydrate and lipid oxidation rates (g/min), respectively, and 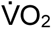 and 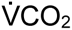 denote oxygen consumption and carbon dioxide production (L/min), respectively.

#### Subjective appetite scores

Subjective appetite scores will be assessed at baseline and at the end of each hour using computerized 100-mm visual analog scales (AVAS, Neurobehavioral Research), measuring desire to eat (“How strong is your desire to eat?” very weak – very strong), hunger (“How hungry do you feel?” not hungry at all – as hungry as I have ever felt), fullness (“How full do you feel?” not full at all – very full), and prospective food consumption (“How much food do you think you could eat?” nothing at all – a large amount) (22). Assessments will be conducted under standardized conditions, free of odours, sounds, and visual stimuli that could influence appetite ratings. Participants will not be permitted to view their previous responses.

#### Serum estradiol and progesterone concentrations

To biochemically verify phase assignment, a baseline venous blood sample will be collected into a BD Vacutainer serum tube (Becton Dickinson), allowed to clot for approximately 30 minutes at room temperature, and then centrifuged at 1,250 RCF for 10 minutes at 4 °C (Centrifuge 5702 R, Eppendorf). The serum will be immediately aliquoted and stored at −80 °C until analysis. Serum β-estradiol and progesterone concentrations will be measured in duplicate by enzyme-linked immunosorbent assay to confirm that hormone concentrations were within the expected physiological ranges for each phase, as previously described by our group (23). If hormone concentrations fall outside the expected physiological ranges for the assigned phase (5th to 95th percentile; early follicular phase: β-estradiol 78–266 pmol/L and progesterone 0.32–1.91 nmol/L; mid-luteal phase: β-estradiol 276–761 pmol/L and progesterone 21–54 nmol/L), the session will be flagged, and the data will be retained but reported separately (9). Primary analyses will be conducted on all participants, with a sensitivity analysis excluding sessions in which phase assignment could not be biochemically confirmed.

### Harms

Adverse events will be monitored throughout each experimental session by research team members who observe participants continuously. Blood pressure and heart rate will be measured in duplicate using a clinical-grade automated sphygmomanometer (UA-651CN, A&D Medical) at baseline and at the end of each hour. Any adverse event or participant complaint will be recorded as it occurs. Given the minimal-risk nature of the protocol, no formal severity grading system will be applied, and no stopping rules will be prespecified.

### Participant timeline

Participation in this trial will involve one preliminary visit and two experimental sessions (Figure 1).

### Sample size

The parent trial was powered to detect differences in plasma triglyceride concentrations. An a priori power analysis was conducted using GLIMMPSE (version 3.1.3) and data from a previous study investigating postprandial plasma triglyceride concentrations in premenopausal females after a high-fat meal during the follicular phase (23). An effect size of 10 % was selected for the present study, acknowledging that a minimal clinically meaningful difference in this context has not been established. Within-participant factors included menstrual cycle phase (early follicular, mid-luteal) and time (0, 30, 60, 90, 120, 180, 240, 300, and 360 minutes; corresponding to the blood sampling schedule in the parent trial). The analysis indicated that a sample size of 10 participants would be sufficient to detect a significant main effect of phase with 80 % power and α = 0.050.

For fasting resting energy expenditure and the thermic effect of food, a second a priori power analysis was conducted using G*Power (version 3.1.9.4) for a repeated-measures analysis of variance within factors, with phase (early follicular, mid-luteal) and time (baseline and end of each hour for 6 hours; 7 measurements) as within-participant factors. An effect size of f = 0.33 was adopted based on the meta-analytic estimate reported by Benton et al. for the effect of menstrual cycle phases on fasting resting energy expenditure, used as a conservative approximation in the absence of a pooled effect size for the thermic effect of food (10). A correlation of 0.5 among repeated measures and a nonsphericity correction of 1 were assumed. The analysis indicated that a sample size of 12 participants would be sufficient to achieve 80 % power at α = 0.05.

Accordingly, a recruitment target of 12 participants with complete datasets was established, as this satisfies the requirements of both power analyses and ensures that the minimum analyzable sample for the primary trial outcome (n = 10) is met even in the event of up to 20 % dropout. Recruitment will continue until 12 participants have completed both experimental sessions. If more than 12 eligible individuals are screened and provide informed consent before data collection is complete (for example, due to the staggered nature of session scheduling across participants), all consented participants who complete both experimental sessions will be included in the analyses. No interim analyses will be conducted or are pre-planned, and no stopping guidelines were prespecified.

### Recruitment

Participants will be recruited from the Ottawa–Gatineau region through convenience sampling. Recruitment will primarily take place within the University of Ottawa community through word of mouth and trial registration. Interested individuals will contact the research team directly and undergo a brief telephone or email pre-screening to assess preliminary eligibility before being invited to the preliminary visit, at which formal eligibility will be confirmed and written informed consent obtained. Recruitment began on 26 March 2026 and is expected to continue until the target sample of 12 participants has been enrolled, with a projected study completion date of 31 August 2026. A flow chart depicting the screening, enrolment, and completion of participants through both experimental sessions will be included in the Stage 2 manuscript in accordance with CONSORT 2025 reporting guidelines (15).

### Randomization and blinding

Randomization will not be performed. Session order will be determined by scheduling convenience (follicular then luteal, or luteal then follicular), as the inherent temporal structure and personal nature of the menstrual cycle preclude randomization of phase assignment. Masking of participants or data collectors to the menstrual cycle phase will not be feasible, given the procedures used to confirm phase assignment. However, analyses of the present secondary outcomes will be performed by K.M., who will be masked to phase assignment; phase labels will be removed from the dataset prior to transfer and will be withheld until all analyses are complete. No unblinding will occur during analysis.

### Data collection methods

All data for primary and secondary outcomes will be collected by S.L. (graduate student at the master’s level) during each experimental session using standardized procedures (as detailed in the *Outcomes* section), with assistance from N.G. (doctoral candidate) and supervision from P.I. (full professor and principal investigator). Primary outcomes will be assessed by S.L. and N.G.; secondary outcomes will be assessed by N.G., N.B. (doctoral candidate), and K.M. (postdoctoral fellow). Retention will be promoted by modest financial compensation for participants who complete experimental sessions. No plan will be in place to record the reason for non-retention (e.g., withdrawal from the trial or loss to follow-up).

### Data management

All de-identified data will be recorded and stored on a password-protected institutional server accessible only to the research team; data will be entered by S.L. and verified by N.G. to reduce transcription errors. Blood samples will be processed and stored at −80 °C in a key-card-protected laboratory. Participant data will be identified by a unique numeric code; no personally identifiable information will be stored alongside outcome data.

### Statistical analysis

Baseline values (0 minutes; fasting) for all variables and the total thermic effect of food over the 360-minute postprandial period will be compared between phases using a paired Student’s t-test. Postprandial outcomes will be analyzed using linear mixed-effects models fitted by restricted maximum likelihood to evaluate the effects of menstrual cycle phase (early follicular, mid-luteal) and time (60, 120, 180, 240, 300, and 360 minutes), both fitted as categorical predictors, on each outcome, with participant identification included as a random effect and the fasting (0 minutes) measurement of each respective outcome included as a continuous covariate to account for intra-individual baseline variability. For the thermic effect of food, which is derived by subtracting fasting resting energy expenditure from each postprandial measurement, session order will be included as a covariate in place of the fasting value to account for potential order effects in the non-randomized crossover design.

Models will be initially fitted with and without a phase × time interaction. The optimal fixed-effect structure will be determined using Akaike’s information criterion (AIC); a difference of two or more AIC units (ΔAIC ≥ 2) in favour of the multiplicative model will be considered evidence of improved fit (24). If the multiplicative model meets this threshold but the interaction term is not statistically significant at α = 0.050, the additive model will be chosen as the final model. Model assumptions (homoscedasticity and normality of residuals and random effects) will be assessed by visual inspection of diagnostic plots. When a significant main effect or interaction is observed, post hoc pairwise comparisons will be performed with Tukey’s Honestly Significant Difference test. All participants who complete both experimental sessions will be included in each analysis. Statistical significance will be set at α = 0.050. Analyses will be performed in *jamovi* (version 2.2.6; software versions reflect those available at the time of protocol writing and may be updated at Stage 2) with the GAMLj3 module (version 3.3.4). Descriptive data will be presented as mean (standard deviation). Differences between menstrual cycle phases and changes over time will be expressed as mean (95 % confidence interval, lower bound to upper bound) derived from estimated marginal means.

Equivalence testing will also be performed using the Two One-Sided Tests procedure in *jamovi* with the TOSTER module (version 0.4.1), applied to the estimated marginal means at the phase level (i.e., a single mean value per phase, collapsed across time points, derived from the baseline-adjusted mixed-effects models). Equivalence bounds will be set at ±0.33 standard deviations for fasting resting energy expenditure and thermic effect of food, corresponding to the smallest effect size the study was powered to detect. For the respiratory exchange ratio, bounds will be set at ±0.02 RER units, corresponding to the within-subject measurement error of the indirect calorimetry system in our laboratory (coefficient of variation = 2.3%; r = 0.99). For subjective appetite scores (desire to eat, hunger, fullness, and prospective food consumption), bounds will be set at ±10 mm on the 100 mm visual analog scale, corresponding to a realistic difference between two meals (25). Equivalence testing will not be applied to carbohydrate and lipid oxidation rates, for which no a priori equivalence bounds can be justified, nor to serum β-estradiol and progesterone concentrations, which serve as biochemical verification measures for phase assignment rather than as experimental outcomes. Notably, however, the present study was powered specifically for fasting resting energy expenditure and the thermic effect of food; all remaining outcomes were not included in the sample size calculation and should, therefore, be interpreted as exploratory, with findings considered hypothesis-generating.

Individual data points will be excluded from analysis if they are identified as technically invalid; for example, if the indirect calorimetry system fails to reach steady state, if a venous blood sample is hemolyzed, or if a visual analog scale response is missing due to equipment failure. No a priori criteria for statistical outlier removal will be applied; all valid data points will be retained regardless of their distance from the group mean. Any excluded data points or sessions will be reported transparently. Missing data will be considered as missing at random and not imputed.

GraphPad Prism (version 11.0) will be used to generate figures presenting raw data as the mean (standard deviation) and individual points. Model-derived estimated marginal means with 95 % confidence intervals will be presented in a supplemental file to visualize covariate-adjusted effects.

### Data monitoring committee and trial monitoring

No data monitoring committee will be established for this trial. Given the minimal-risk nature of the procedures, the short duration of each experimental session, the absence of planned interim analyses, and the small sample size, formal independent data monitoring will not be considered necessary. Oversight of trial conduct will be provided by the principal investigator (P.I.). Trial conduct will be monitored informally by the principal investigator on an ongoing basis. Given that all experimental sessions will be conducted at a single site by the same research team under direct supervision, no formal remote or on-site monitoring plan was developed. No independent monitoring body or external auditor will be involved.

## DATA AVAILABILITY STATEMENT

De-identified participant data for this study will be available upon reasonable request to the corresponding author and with a signed access agreement, in accordance with the conditions of ethics approval granted for this trial by the Research Ethics Board of the University of Ottawa. Analysis code generated in *jamovi* will be made available as a supplementary file accompanying the Stage 2 manuscript. The questionnaires that will be used in this study are described in full in their respective original validation publications, which are cited in the Methods section.

## AUTHOR CONTRIBUTIONS

All those designated as authors meet all four criteria for authorship outlined by the International Committee of Medical Journal Editors, and all who meet the four criteria are identified as authors. All authors approved the final version of the manuscript; agree to be accountable for all aspects of the work in ensuring that questions related to the accuracy or integrity of any part of the work are appropriately investigated and resolved; and all persons designated as authors qualify for authorship, and all those who qualify for authorship are listed. **Nicholas Goulet:** Conceptualization, Methodology, Project Administration, Investigation, Data Curation, Formal Analysis, Writing – Original Draft, Writing – Review & Editing, Visualization. **Sydney Lyndon:** Methodology, Investigation, Data Curation, Writing – Review & Editing. **Noémie Beauregard:** Formal Analysis, Writing – Review & Editing. **Kurt McInnis:** Formal Analysis, Writing – Review & Editing. **Jean-François Mauger:** Validation, Writing – Review & Editing. **Éric Doucet:** Conceptualization, Writing – Review & Editing. **Pascal Imbeault:** Conceptualization, Project Administration, Writing – Review & Editing, Supervision, Funding Acquisition.

## DISSEMINATION PLAN

Trial results will be submitted for publication in a diamond open-access journal. All authors will review and approve the manuscript prior to submission.

## Acknowledgement

All authors have read and approved this version of the manuscript. This article was last modified on 8 September 2026.

